# COVID-SGIS: A smart tool for dynamic monitoring and temporal forecasting of Covid-19

**DOI:** 10.1101/2020.05.30.20117945

**Authors:** Clarisse Lins de Lima, Cecilia Cordeiro da Silva, Ana Clara Gomes da Silva, Eduardo Luiz Silva, Gabriel Souza Marques, Lucas Job Brito de Araújo, Luiz Antônio Albuquerque Júnior, Samuel Barbosa Jatobá de Souza, Maíra Araújo de Santana, Juliana Carneiro Gomes, Valter Augusto de Freitas Barbosa, Anwar Musah, Patty Kostkova, Wellington Pinheiro dos Santos, Abel Guilhermino da Silva Filho

## Abstract

**Objective:** The new kind of coronavirus SARS-Cov2 spread to countries in all continents in the World. The coronavirus disease 2019 (Covid-19) causes fever, cough, sore throat, and in severe cases shortness of breath and death. To evaluate strategies, it is necessary to forecast the number of cases and deaths, in order to aid the stakeholders in the process of making decisions against the disease. We propose a system for real-time forecast of the cumulative cases of Covid-19 in Brazil.

**Study Design:** Monitoring of all Brazilian cities using oficial information from the National Notification System, from March to May 2020, concentrated on Brazil.io databases. Training and evaluation of ARIMA and other machine learning algorithms for temporal forecasting using correlation indexes (Pearson’s, Spearman’s, and Kendall’s) and RMSE(%). Validation from the relative errors of the following six days.

**Methods:** Our developed software, COVID-SGIS, captures information from the 26 states and the Distrito Federal at the Brazil.io database. From these data, ARIMA models are created for the accumulation of confirmed cases and death cases by Covid-19. Finally, six-day forecasts graphs are available for Brazil and for each of its federative units, separately, with a 95% CI. In addition to these predictions, the worst and best scenarios are also presented.

**Results:** ARIMA models were generated for Brazil and its 27 federative units. The states of Bahia, Maranhão, Piauí, Rio Grande do Norte and Amapá, Rondônia every day of the predictions were in the projection interval. The same happened to the states of Espírito Santo, Minas Gerais, Paraná and Santa Catarina. In Brazil, the percentage error between the predicted values and the actual values varied between 2.56% and 6.50%. For the days when the forecasts outside the prediction interval, the percentage errors in relation to the worst case scenario were below 5%. The states of Bahia, Maranhão, Piauí, Rio Grande do Norte, Amapá, and Rondônia every day of the predictions were in the projection interval. The same happened to the states of Espírito Santo, Minas Gerais, Paraná and Santa Catarina.

**Conclusion:** The proposed method for dynamic forecasting may be used to guide social policies and plan direct interventions in a robust, flexible and fast way. Since it is based on information from multiple databases, it can be adapted to the different realities, becoming an important tool to guide the course of politics and action against Covid-19 pandemic worldwide.

## 1. Introdução

The world are facing a pandemic caused by the new coronavirus SARS-Cov2. The probable origin of the SARS-Cov2 is from a pangolin’s virus (Zhang et al., 2020). SARS-Cov2 has a high transmission rate. During later December 2019 and May 2020 more than 4.7 million people were infected in 216 countries. (WHO, 2020). Some coronavirus disease 2019 (Covid-19) symptoms are fever, cough, sore throat and gastrointestinal infection (Guo et al., 2020). But in severe cases Covid-19 can lead to shortness of breath and death. Until later May 317 thousands people died due Covid-19 (WHO, 2020).

The gold standard test to diagnosis Covid-19 is the Reverse Transcription Polymerase Chain Reaction (RT-PCR) with DNA sequencing and identification (Döhla et al., 2020). Nevertheless, RT-PCR needs several hours to give a result (Döhla et al., 2020). While RT-PCR identify directly the virus presence, in other hand, rapid test are nonspecific. They detect the serological evidence of recent infection like antibodies. However the production of antibodies starts after some days or even weeks after the infection (Okba et al., 2020). Besides that, this kind of tests could recognize antigens of other viruses, like flu and other coronaviruses (Li et al., 2020). What compromise the test accuracy. For example, tests based on IgM/IgG antibodies realized in samples collected in the first week of illness have 18.8% of sensitivity and 77.8% of specificity (Liu et al., 2020a).

The best strategy to decrease the transmission rate of the SARS-Cov2 is the social isolation and quarantine. Its main goal is to provide a safe physical distance between infected and uninfected people. Chen et al. (2020) reported that measure as cancelled or postponed large public events and closed non-essential activities have helped to reduce the spread of the virus in China. Social isolation and quarantine also prevent that asymptomatic infected people spread the disease (Chen et al., 2020; Day, 2020). Other works also show good results in reduce the virus transmission with social isolation in Italy Day (2020), Switzerland Salathé et al. (2020) and Brazil (Oliveira et al., 2020).

One of the approaches used to combat diseases is the prediction of the number of cases based on the behavior of past events. In the case of arboviruses, for example, the time series of the number of cases and climatic variables are used to predict future behaviors (Guo et al., 2017; Siriyasatien et al., 2018; Baquero et al., 2018). Considering the SARS-Cov2, predicting the pandemic trends could be crucial to avoid the virus spreading. Because of that many models basing in mathematical model as Susceptible-Infected-Removed (SIR) and its variations have being used to forecast Covid-19 pandemic trends (Wangping et al., 2020; Bastos and Cajueiro, 2020; Liu et al., 2020b; Anastassopoulou et al., 2020; Yang et al., 2020; Ardabili et al., 2020).

The main goal of this work is to present a methodology for monitoring and forecasting of cases and deaths of Covid-19 in Brazil and its federative units. The system is basing in a web application that use information from multiple databases. This informations are used for forecasting in national and state level. We used an ARIMA method (Hyndman and Khandakar, 2008) adapted to a very dynamic disease as Covid-19. Our model is basing in a dynamic forecasting because the model is training each day with a maximum window of three days. Thus our model is flexible and robust considering that it’s adapt each day. That makes policy measures been perceived by the method. Besides that, the model gives three possible scenarios: the expected case with 95% confidence interval (CI) and also the worst and best cases.

This paper is organized as follows: Section 2 we discuss about studies that used mathematical models to predict trends of Covid-19 pandemic. Section 3 we describe our proposed method and it also reviews the theoretical concepts that concern this work. In section 4 we present and analyze the results. Section 5 draws discussions and the work finalize in Section 6 with the conclusions.

## 2. Related Works

Several Covid-19 prediction models have emerged around the world. Because of the many uncertainties surrounding the behavior of the virus, these models have guided the development of public health strategies and the application of policies that promote social isolation. Given this scenario, several studies have sought to adapt conventional epidemiological models to predict this pandemic trend.

Wangping et al. (2020), for instance, used an extended Susceptible-Infected-Removed (eSIR) model to predict epidemics trends in Italy and compared with Hunan, China due to its similarity of population size with Italy. eSIR model is a version of the Susceptible-Infected-Removed (SIR) classic model. SIR model takes three differents classes: susceptible, infected, removed (recovered and dead). This model was presented by Kermack and McKendrick in their article published in 1927 (Kermack and McKendrick, 1927), by Britton in their book entitled “Essential Mathematical Biology” (Britton, 2003) and by Brauer et al. in the book “Mathematical Epidemiology’ (Brauer et al., 1945). SIR model is used to investigate diseases as measles and chickenpox. This model is represented by the following equations:

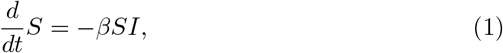

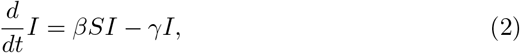

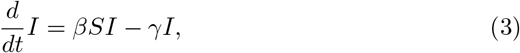

where S(t) is the population susceptible individuals, I(t) is the symptomatic infected individuals, R(t) is the recovered individuals with immunity, *β* is the contact rate between susceptible and infected individuals, and *γ* is the transfer rate from I to R. The diagram in the Figure 1 explains the SIR model.

**Figure 1:**
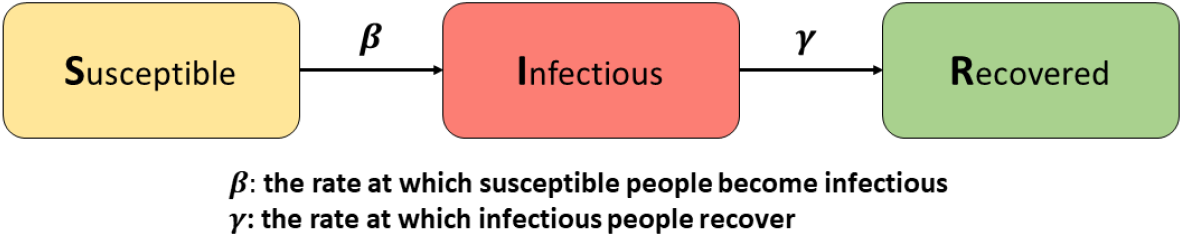
Flow chart of the SIR model. The classic SIR model considers three classes: Susceptible, Infectious and Recovered. *β* means the rate at which susceptible people can become infectious, and *γ* is the rate at which infectous people recover.

Whereas SIR model uses a constant transmission rate eSIR can adapt the time-varying transmission rate in the population and also it uses time-varying isolation measures. Using eSIR Wangping et al. (2020) predict that the reproduction number, R0, for Covid-19 was respectively 3.15 (95% CI: 1.71–5.21) in Hunan and 4.10 (95% CI: 2.15–6.77) in Italy. Due to the discrepancy between the values they concluded that this needs to be confirmed by further studies.

In a similar way, Bastos and Cajueiro (2020) developed models based on SIR. Their main objective was to predict the evolution of the pandemic in Brazil through data analysis in the period from February 25, 2020 to March 30, 2020. Data were provided by the Ministry of Healthy of Brazil. For this, the authors modified the SIR model, creating two other versions: SID (Susceptible-Infected-Dead) and SIASD (Susceptible-Infected-Asymptomatic-Symptomatic-Dead). In the first version, the authors modified the original model in order to estimate the percentage of people who die from Covid-19. This new parameter also changes the total number of individuals in the population. On the other hand, the second version seeks to improve the SID model. It considers that a relevant part of the population is infected, but asymptomatic. Then, in the SIASD model, the variable of infected people is divided into two groups: symptomatic and asymptomatic. In addition, the authors modified the transmission factor taking into account the effect of social distance policies adopted during the selected period. The addition of this parameter allows the model to evaluate the effectiveness of the measures adopted. In order to estimate the parameters of the models, the work sought to minimize the quadratic error between the real data and the estimated values. The curves were constructed with a 95% confidence interval. The authors also considered the underreporting of cases, due to the lack of tests and the government’s recommendation to test only patients with severe symptoms. Finally, both models indicated that social distance policies were able to minimize contagion. It was also possible to conclude that policies adopted for a short period are only able to postpone the peak of the pandemic. Thus, the authors indicated the dates considered optimal to end the quarantine period, in order to reduce the number of infections. In particular, the SIASD model predicted a greater number of infections than the SID model. It also indicated a lower peak for symptomatic patients, who are those who need medical attention.

In the study from Liu et al. (2020b), they also proposed a modified SIR model to predict the cumulative number of cases of Covid-19 in China. Their model was based on early data and is defined by the following system of differential equations and the flowchart in Figure 2.

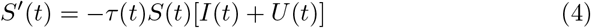

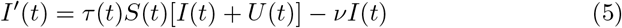

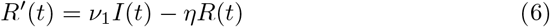

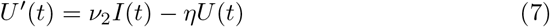

where *S*(*t*) is the number of individuals susceptible to infection at time t, *I*(*t*) is the number of asymptomatic infectious individuals at time t, *R*(*t*) is the number of reported symptomatic infectious individuals at time t, *U*(*t*) is the number of unreported symptomatic infectious individuals at time t, *τ*(*t*) is the transmission rate at time t, 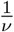 is the average time during which asymptomatic infectious are asymptomatic, and 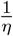 is the average time symptomatic infectious have symptoms.

**Figure 2:**
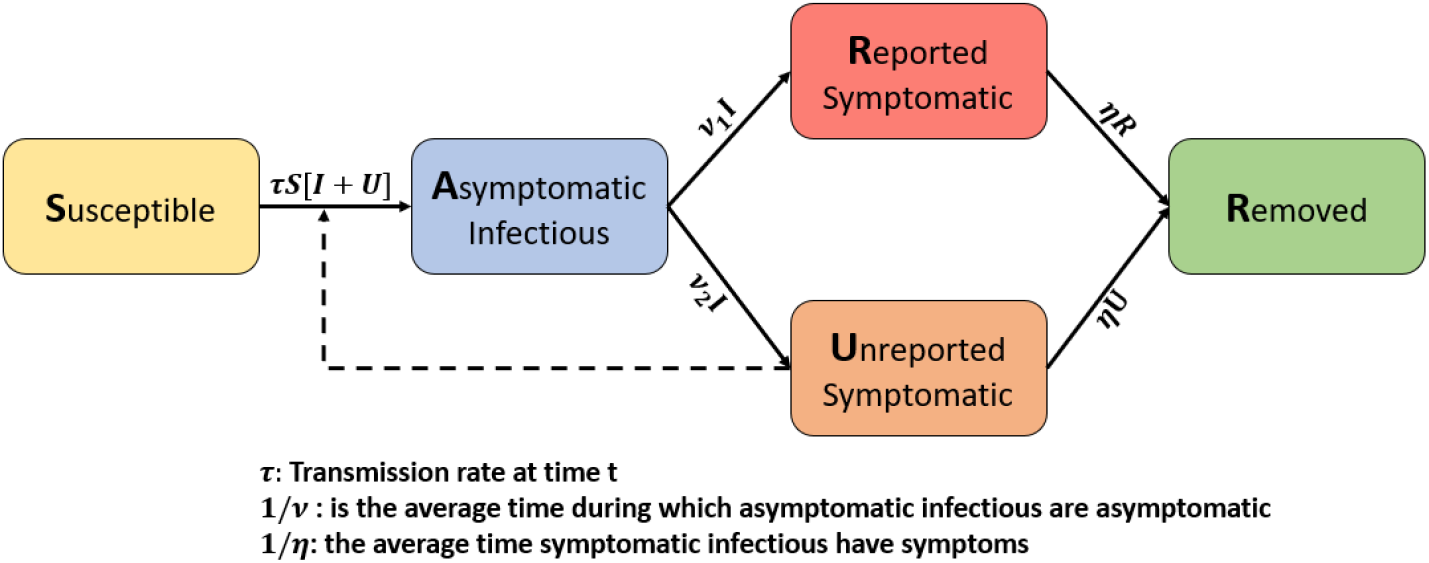
Model flow chart adapted from Liu et al. (2020b). They proposed a modified SIR model by adding asymptomatic infectous people and by dividing symptomatic cases into two classes: reported and unreported.

This model provides information regarding to the number of both asymptomatic and symptomatic individuals. It also estimates the amount of reported and unreported cases from the symptomatic group. From this study, the authors found that the estimation of unreported cases is extremely important to understand the severity of Covid-19. They also showed the importance of considering the asymptomatic infectious cases to estimate disease transmission rate. From their results, the authors demonstrated the importance of implementing severe control measures by the government to decrease transmission.

Anastassopoulou et al. (2020) proposed a Susceptible-Infectious-Recovered-Dead (SIRD) model to forecast the basic reproduction number (*R*_0_) of SARSCov2 and the daily rates of infection mortality and recovery. *R*_0_ measures the average number of secondary cases that result from a single infectious case. From the reproduction number, a system may predict the spreadability of a infectious disease.

This approach was based on the daily available data of new confirmed cases in Hubei province, China, and is described by the following equations:

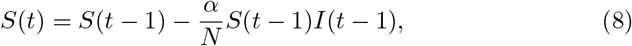

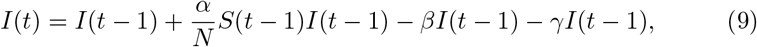

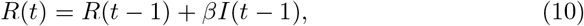

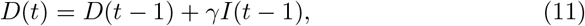

where *S*(*t*) is the number of susceptible individuals at time t, *I*(*t*) is the number of infected individuals at time t, *R*(*t*) is the number of recovered individuals at time t, *D*(*t*) is the number of dead individuals at time t, *α* is the estimation of the infection rate, *β* is the estimation of the recovery rate, *γ* is the estimation of the mortality rate, and *N* is the population size.

This model fits the behavior of Covid-19 in Hubei, being able to estimate key epidemiological parameters. The approach was also successful in modelling and forecasting the spread of Covid-19 in China. Nevertheless, their SIRD model did not consider important factors that changes disease dynamics, such as the effect of the incubation period, the characteristics of the population and effect of restritive policies. The authors also point out to the urgent need for testing and available official data. These factors are crucial to build accurate forecast models, since they are the key way to tune the model by providing a more realistic amount of infected cases.

On the other hand, Yang et al. (2020) proposed a modified Susceptible-Exposed-Infectious-Removed (SEIR) model and the Artificial Intelligence method Long-Short-Term-Memory (LSTM), to predict epidemics trend of Covid-19 in China under public health interventions. SEIR model is another conventional prediction model, which is composed by four individual classes: Susceptible, Exposed to the virus or in the latent period, Infected and Removed. This model was first introduced by Kermack and McKendrick in 1927 (Kermack and McKendrick, 1927) and is used to understand illnesses like flu for example. SEIR model uses the following equation system:

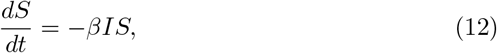

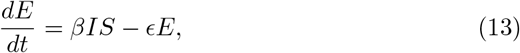

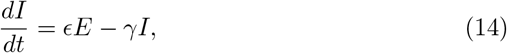

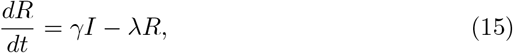

where S(t) is the population susceptible individuals, E(t) represents the individuals exposed to the disease or in a latent period, I(t) is the symptomatic infected individuals, R(t) is the recovered individuals with immunity, *β* is the contact rate between susceptible and infected individuals, *ε* is the transfer rate from class S to E, and *γ* is the transfer rate from I to R. The diagram in the Figure 3 explains the SEIR model.

**Figure 3:**
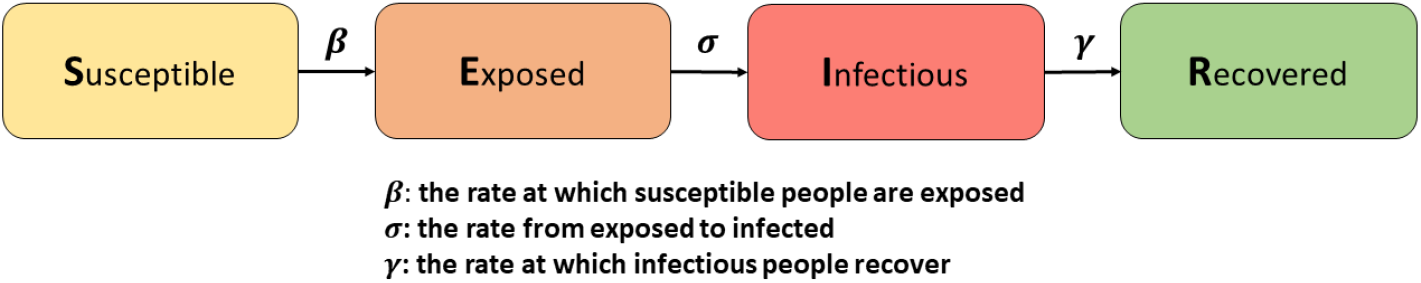
Flow chart of the SEIR model. SEIR model is another conventional prediction model. It is composed by four individual classes: Susceptible, Exposed to the virus, Infected and Removed. In contrast to the SIR model, *β* is the rate at which susceptible people become exposed, *ε* is the rate from exposed to infected, and *γ* is the rate at which infectious people recover.

Yang et al. (2020) modified the SEIR model by incluiding the parameters: move-in, move-out and the contact rate before and after the implementation of control policies. They also used the recurrent neural network Long-Short-Term-Memory (LSTM) to corroborate their model predictions. LSTM was trained on the 2003 SARS epidemic statistics, with available cases between April and June of 2003. They predicted that the disease in China will peak in late February and end in late April by a combination of their methods.

In contrast to previous works, Ardabili et al. (2020) made a comparative analysis of machine learning techniques for predicting the coronavirus outbreak. The authors argue that more traditional models, such as SIR and SEIR, are insufficient to model the pandemic. The reasons are the quarantine and social distance policies applied by many governments in an iterative way, and the lack of data that reveals the real scenario, since the reported data do not actually correspond to the number of infected people. These factors impose great limits on the generalization ability of these conventional models. Thus, the work explores ML techniques to find the best model that estimates time-series data. Data were collected on the worldometers website in five countries: Italy, Germany, Iran, USA and China, corresponding to a period of 30 days. Initially, simple mathematical models were tested (Logistic, Logarithmic, Quaddratic, Compound, Power and exponential). In order to estimate the parameters, three optimization algorithms were tested: Genetic Algorithm (GA), Particle Swarm Optimization (PSO), and the Gray Wolf (GWO) optimizer. Considering the processing time, the RMSE and the correlation coefficient as evaluation metrics, the GWO optimizer presented the best results, while the logistic model showed the smallest errors in the prediction of the Covid-19 outbreak for the five countries. However, these models showed low accuracy and low generalization capacity, being unable to extrapolate the data beyond 30 days. In the following, two ML methods were introduced for time-series prediction: Multi-Layer Perceptron (MLP) and adaptive network-based fuzzy inference system (ANFIS). Both were tested in two different scenarios: Scenario 1 with data processed weekly, and Scenario 2, with daily sampling of consecutive days. In the case of MLP, they tested 8, 12 and 16 neurons. For ANFIS, the membership function types were Tri, Trap and Gauss. The comparison between analytical and intelligent models indicated that MLP presented better results in both scenarios, being able to achieve a good approximation with the real data. Longterm data (up to 150 days) were also extrapolated, which reported an outbreak progression. Finally, the authors concluded that ML techniques can be useful in predicting the Covid-19 pandemic, being able to overcome challenges found in traditional models.

## 3. Materials and Methods

### 3.1. Proposed Method

In this work, we proposed a system for real-time forecast of the cumulative cases of Covid-19 in Brazil and its 27 federative units. The system operates as follows: Each Brazilian state feeds a database of Covid-19 notifications. All of this information is gathered in a general database, the Brazil.io. Then, our developed software, the COVID-SGIS, captures Covid-19 data daily made available by the Brasil.io portal^1^ through the SGIS web crawler. This accumulated data is organized in a CSV file. From these data, training sets are formed and the models are created for the accumulation of confirmed cases and death cases by Covid-19. With the generated models, 6-day forecasts are made with a 95% confidence interval which are available to the user. The forecast graphs of the accumulated confirmed cases and deaths by Covid-19 are available for Brazil and for each of its federative units, separately. In addition to the predictions, the worst and best scenarios are presented for the number of confirmed cases and the number of deaths by Covid-19. The Figure 4 shows the general schematic of this solution.

**Figure 4:**
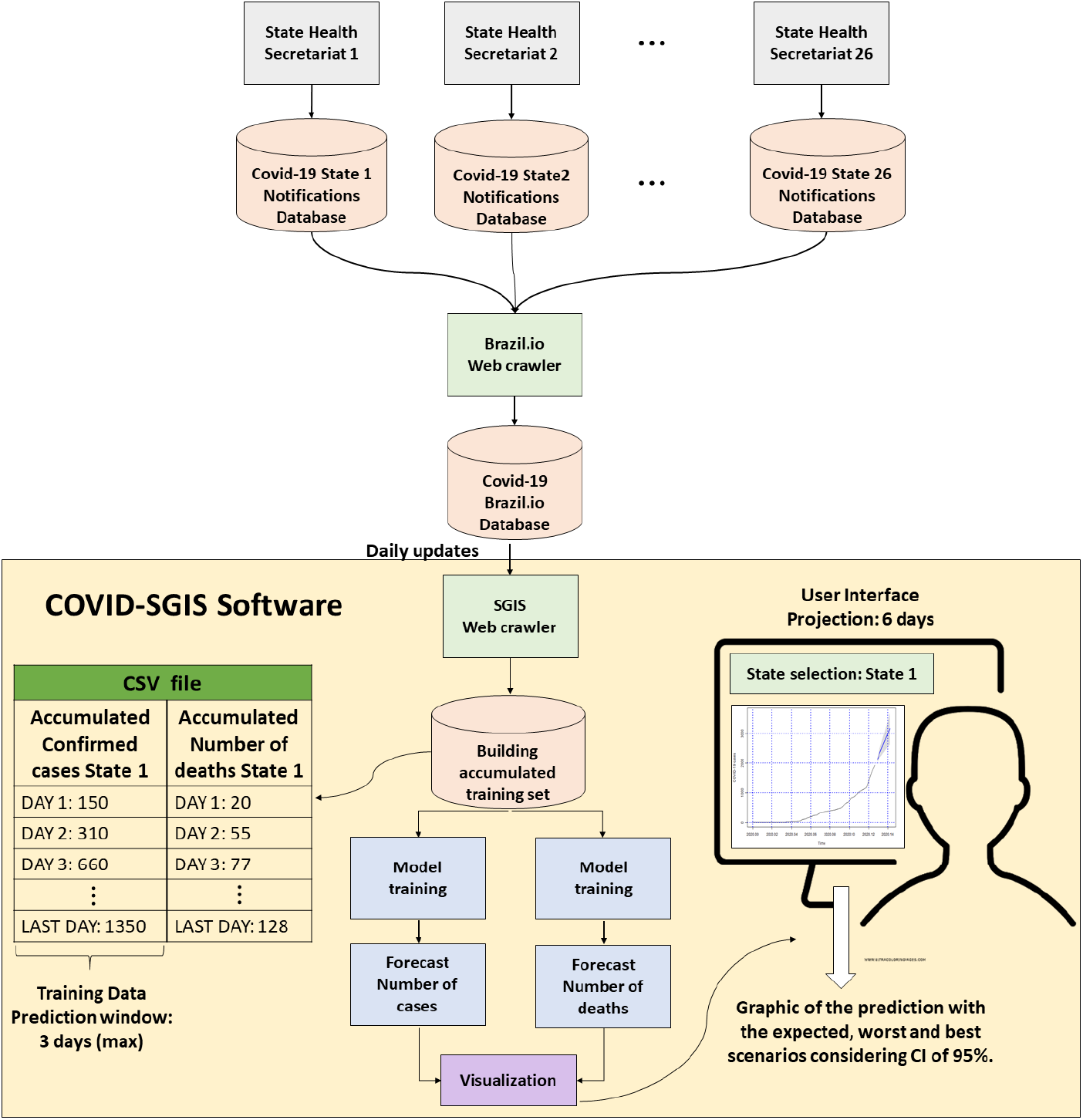
Proposed method: Each of the Health Secretaruat of the 26 Brazilian states plus the Distrito Federal (the autonomous disctrict in which is inserted the national capital) is responsible for feeding a notification base. All of this information is available on Brazil.io. Our Covid-SGIS software is updated daily with data from Brazil.io. A file in CSV format is organized with the accumulated data. From them, training sets of the model can be formed. After the ARIMA model training, the user can view the prediction of the number of cases and deaths for each of the states, with a 6-day projection.

### 3.2. Confirmed cases database

We used the data referring to confirmed cases available in the Brasil.io portal^2^ in our temporal prediction approach. Brasil.io portal provides the records of confirmed cases and deaths obtained through the bulletins of the State Health Secretariats.

In this work, we selected data related to confirmed cases of Covid-19 for all Brazil’ federative units. For each federative unit the records is from the first date of confirmation of the disease until the last update on May 5, 2020.

### 3.3. ARIMA model

ARIMA models are classic models widely used to predict future behaviors of stationary time series. ARIMA’s acronym indicates the combination of autoregressive (AR) and moving average (MA) models. The “I” indicates the model for the original undifferenced series, which was differenced *d* times until it became a stationary series before fitting the ARMA(p,q) process. In the ARIMA(p, d, q) model, *p* and *q* correspond to the orders of the AR and MA models, respectively, while the *d* corresponds to the level of differencing. The equation 16 represents the mathematical expression of the model, where *y_t_* is the differenced series, *c* and *φ* are the parameters of the model and *ε* is the random error in time *t* (Chakraborty and Gosh, 2020; Hyndman and Athanasopoulos, 2018).

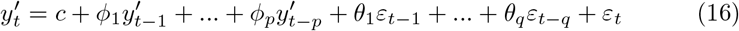

The construction of the ARIMA model (p, d, q) occurs according to the following steps (Chakraborty and Gosh, 2020; Hyndman and Athanasopoulos, 2018):

1. Evaluate the stationarity of the series (if it is not stationary, the differencing is applied util the stationarity is achieved);
2. Estimate the *p* and *q* parameters based on the autocorelation function (ACF) and the partial autocorrelation function (PACF) plots;.
3. Evaluate the best fitted forecasting model using Akaike Information Criterion (AIC) and the Baysian Information Criterion (BIC).

The function *auto.arima()* in the forecast package in R, automatically calculates the *p*, *d* and *q* parameters and returns a fitted model. The *forecast()* function of this same package can be used to predict based on the model adjusted by the *auto.arima()* function (Hyndman and Khandakar, 2008).

In this context, from the data collected regarding the accumulated number of confirmed cases by state, we created a database corresponding to the historical series of accumulated confirmed cases of Covid-19 for Brazil and each of its federal units. We considered the period from the first notification date of the disease until May 5, 2020. From this historical series of accumulated cases of Covid-19, we built ARIMA models for each state, and for the Distrito Federal, and for Brazil. These models were generated using the function *auto.arima()* of R tool, version 3.6.3 ^3^. Each model built was used to carry out the projection of the accumulated cases of the disease for 6 days, with a confidence interval of 95%. In order to evaluate the performance of the forecasts, we used accumulated data from May 6 to 11, which were updated on May 13.

### 3.4. Metrics

We selected two metrics to evaluate the models: the correlation coefficient and the Relative Quadratic Error (RMSE percentage). The correlation coefficient is a statistical measure between expected and predicted values. This value varies from −1 to 1.. When it approaches 1, it indicates a strong positive correlation. When the correlation coefficient is close to −1, it indicates that the variables have a strong negative correlation. And when the correlation coefficient is close to zero, it indicates that there is no correlation between the variables (Witten and Frank, 2005). The value of the correlation coefficient serves as global evaluator of the model, so it is possible to obtain a high correlation coefficient and at the same time obtain high values for local errors. For this reason, it can not be the only metric for assessing model performance. In order to avoid a superficial evaluation of the regressors, whe chose the relative quadratic error as an evaluation metric. The Equation 17 shows the expression of the calculation of the relative quadratic error, where *p* is the predicted value and *a* is the actual value.

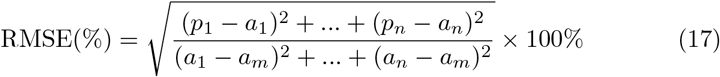

## 4. Results

### 4.1. ARIMA forecasting

The models built were evaluated taking into account, as global quality, the correlation coefficients of pearson, spearman and kendall. The relative quadratic error (RMSE%) was used with a local quality metric. In this work, a high correlation coefficient is considered to be above 0.9 and a low relative quadratic error to be below 5%. The table 1 shows the evaluation metrics of the results for the models using ARIMA.

**Table 1:** Results of the correlation coefficients of pearson, spearman and kendall, and of the RMSE% for the ARIMA models built for Brazil and its 27 federative units.

| state | model | pearson | spearman | kendall | RMSE% | number of observations |
| --- | --- | --- | --- | --- | --- | --- |
| Acre | ARIMA(2,2,0) | 0.9989 | 0.9951 | 0.9665 | 4.78 | 50.0 |
| Alagoas | ARIMA(0,2,1) | 0.9987 | 0.9937 | 0.9634 | 5.13 | 59.0 |
| Amazonas | ARIMA(0,2,1) | 0.9993 | 0.9990 | 0.9919 | 4.10 | 54.0 |
| Amapá | ARIMA(0,2,1) | 0.9965 | 0.9983 | 0.9833 | 8.78 | 47.0 |
| Bahia | ARIMA(2,2,1) | 0.9997 | 0.9989 | 0.9880 | 2.56 | 61.0 |
| Ceará | ARIMA(0,2,1) | 0.9941 | 1.0000 | 1.0000 | 11.31 | 51.0 |
| Distrito Federal | ARIMA(0,2,1) | 0.9992 | 0.9994 | 0.9915 | 4.02 | 60.0 |
| Espírito Santo | ARIMA(0,2,1) | 0.9987 | 0.9987 | 0.9856 | 5.09 | 62.0 |
| Goiás | ARIMA(0,2,1) | 0.9988 | 0.9997 | 0.9960 | 4.91 | 55.0 |
| Maranhão | ARIMA(0,2,0) | 0.9991 | 0.9987 | 0.9884 | 4.42 | 47.0 |
| Minas Gerais | ARIMA(1,2,2) | 0.9994 | 0.9998 | 0.9977 | 3.46 | 59.0 |
| Mato Grosso do Sul | ARIMA(0,2,1) | 0.9986 | 0.9997 | 0.9960 | 5.40 | 53.0 |
| Mato Grosso | ARIMA(0,2,1) | 0.9983 | 0.9984 | 0.9865 | 6.03 | 47.0 |
| Pará | ARIMA(0,2,1) | 0.9987 | 0.9994 | 0.9919 | 5.52 | 49.0 |
| Paraíba | ARIMA(3,2,0) | 0.9993 | 0.9959 | 0.9758 | 3.91 | 55.0 |
| Pernambuco | ARIMA(0,2,1) | 0.9995 | 0.9995 | 0.9936 | 3.28 | 55.0 |
| Piauí | ARIMA(1,2,0) | 0.9993 | 0.9981 | 0.9827 | 4.00 | 48.0 |
| Paraná | ARIMA(0,2,2) | 0.9994 | 0.9995 | 0.9939 | 3.47 | 55.0 |
| Rio de Janeiro | ARIMA(0,2,2) | 0.9996 | 0.9992 | 0.9926 | 2.96 | 62.0 |
| Rio Grande do Norte | ARIMA(0,2,1) | 0.9974 | 0.9973 | 0.9766 | 7.37 | 55.0 |
| Rondônia | ARIMA(0,2,0) | 0.9982 | 0.9948 | 0.9663 | 6.20 | 47.0 |
| Roraima | ARIMA(0,2,0) | 0.9792 | 0.9932 | 0.9477 | 21.08 | 46.0 |
| Rio Grande do Sul | ARIMA(2,2,2) | 0.9988 | 0.9994 | 0.9928 | 5.10 | 57.0 |
| Santa Catarina | ARIMA(0,2,1) | 0.9925 | 0.9995 | 0.9956 | 12.41 | 55.0 |
| Sergipe | ARIMA(3,2,0) | 0.9960 | 0.9963 | 0.9704 | 9.11 | 53.0 |
| São Paulo | ARIMA(0,2,2) | 0.9992 | 0.9991 | 0.9869 | 4.14 | 71.0 |
| Tocantins | ARIMA(0,2,2) | 0.9979 | 0.9931 | 0.9577 | 6.76 | 49.0 |
| Brasil | ARIMA(0,2,1) | 0.9998 | 0.9972 | 0.9796 | 2.20 | 71.0 |

Tables 2-6, present the results for the forescats using the ARIMA models for Brazil and each of its 27 federative units. The performance of the models was evaluated taking into account the percentage error^*^ between the predicted values and the actual cases of the accumulated cases of Covid-19 and the absolute error between the actual cases and the lower or upper limit of the forecast. The values highlighted in red represent the situations in which the value of the actual cases is outside of the forcast interval and the absolute error** in between the lower or upper limit is greater than 5%.

**Table 2:**
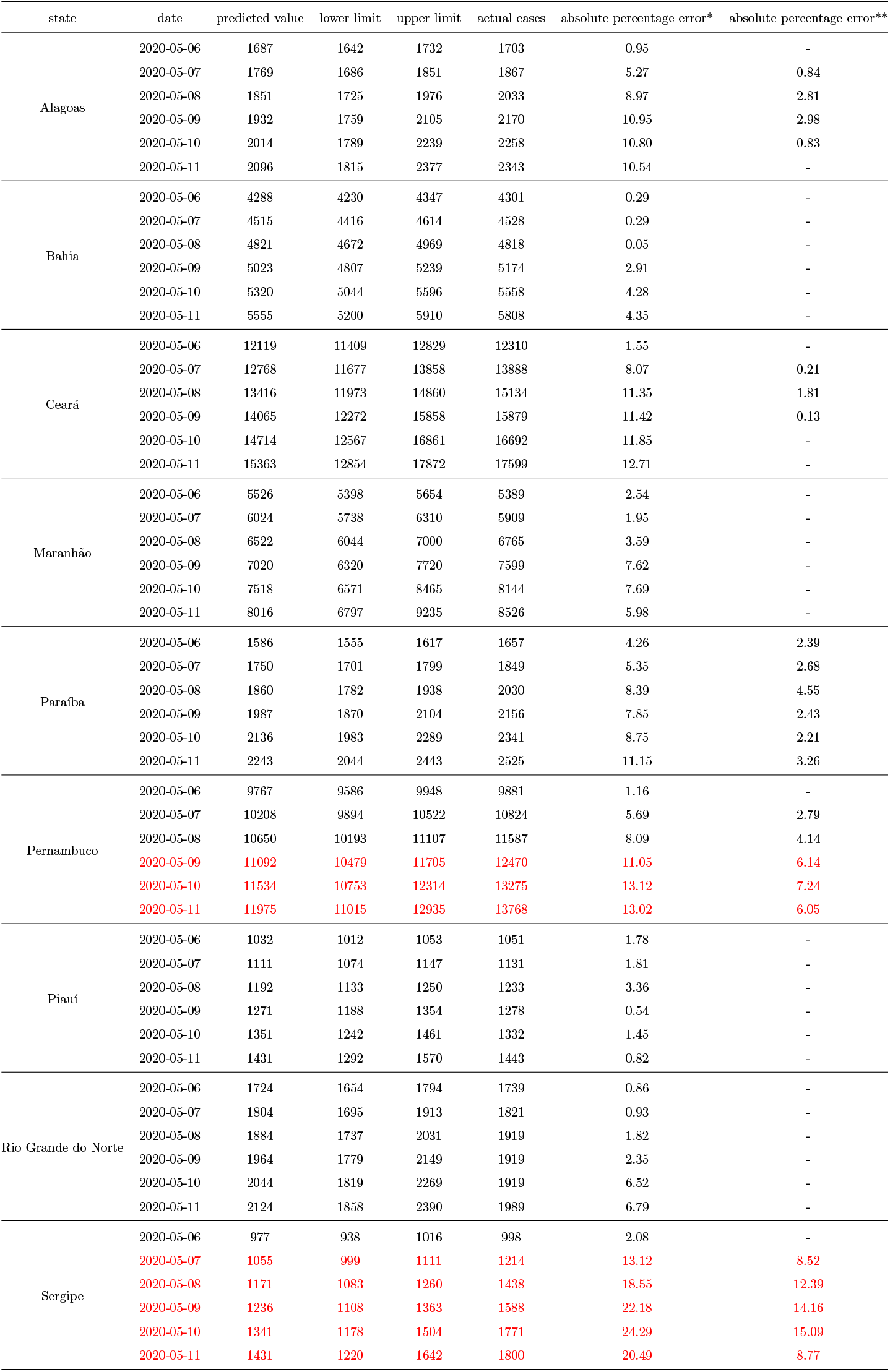
Results of projections of confirmed cases of Covid-19 between May 6 and 11, 2020 for the states of the Northeast Region.

| state | date | predicted value | lower limit | upper limit | actual cases | absolute percentage error* | absolute percentage error** |
| --- | --- | --- | --- | --- | --- | --- | --- |
| Alagoas | 2020-05-06 | 1687 | 1642 | 1732 | 1703 | 0.95 | - |
|  | 2020-05-07 | 1769 | 1686 | 1851 | 1867 | 5.27 | 0.84 |
|  | 2020-05-08 | 1851 | 1725 | 1976 | 2033 | 8.97 | 2.81 |
|  | 2020-05-09 | 1932 | 1759 | 2105 | 2170 | 10.95 | 2.98 |
|  | 2020-05-10 | 2014 | 1789 | 2239 | 2258 | 10.80 | 0.83 |
|  | 2020-05-11 | 2096 | 1815 | 2377 | 2343 | 10.54 | - |
| Bahia | 2020-05-06 | 4288 | 4230 | 4347 | 4301 | 0.29 | - |
|  | 2020-05-07 | 4515 | 4416 | 4614 | 4528 | 0.29 | - |
|  | 2020-05-08 | 4821 | 4672 | 4969 | 4818 | 0.05 | - |
|  | 2020-05-09 | 5023 | 4807 | 5239 | 5174 | 2.91 | - |
|  | 2020-05-10 | 5320 | 5044 | 5596 | 5558 | 4.28 | - |
|  | 2020-05-11 | 5555 | 5200 | 5910 | 5808 | 4.35 | - |
| Ceará | 2020-05-06 | 12119 | 11409 | 12829 | 12310 | 1.55 | - |
|  | 2020-05-07 | 12768 | 11677 | 13858 | 13888 | 8.07 | 0.21 |
|  | 2020-05-08 | 13416 | 11973 | 14860 | 15134 | 11.35 | 1.81 |
|  | 2020-05-09 | 14065 | 12272 | 15858 | 15879 | 11.42 | 0.13 |
|  | 2020-05-10 | 14714 | 12567 | 16861 | 16692 | 11.85 | - |
|  | 2020-05-11 | 15363 | 12854 | 17872 | 17599 | 12.71 | - |
| Maranhão | 2020-05-06 | 5526 | 5398 | 5654 | 5389 | 2.54 | - |
|  | 2020-05-07 | 6024 | 5738 | 6310 | 5909 | 1.95 | - |
|  | 2020-05-08 | 6522 | 6044 | 7000 | 6765 | 3.59 | - |
|  | 2020-05-09 | 7020 | 6320 | 7720 | 7599 | 7.62 | - |
|  | 2020-05-10 | 7518 | 6571 | 8465 | 8144 | 7.69 | - |
|  | 2020-05-11 | 8016 | 6797 | 9235 | 8526 | 5.98 | - |
| Paraíba | 2020-05-06 | 1586 | 1555 | 1617 | 1657 | 4.26 | 2.39 |
|  | 2020-05-07 | 1750 | 1701 | 1799 | 1849 | 5.35 | 2.68 |
|  | 2020-05-08 | 1860 | 1782 | 1938 | 2030 | 8.39 | 4.55 |
|  | 2020-05-09 | 1987 | 1870 | 2104 | 2156 | 7.85 | 2.43 |
|  | 2020-05-10 | 2136 | 1983 | 2289 | 2341 | 8.75 | 2.21 |
|  | 2020-05-11 | 2243 | 2044 | 2443 | 2525 | 11.15 | 3.26 |
| Pernambuco | 2020-05-06 | 9767 | 9586 | 9948 | 9881 | 1.16 | - |
|  | 2020-05-07 | 10208 | 9894 | 10522 | 10824 | 5.69 | 2.79 |
|  | 2020-05-08 | 10650 | 10193 | 11107 | 11587 | 8.09 | 4.14 |
|  | 2020-05-09 | 11092 | 10479 | 11705 | 12470 | 11.05 | 6.14 |
|  | 2020-05-10 | 11534 | 10753 | 12314 | 13275 | 13.12 | 7.24 |
|  | 2020-05-11 | 11975 | 11015 | 12935 | 13768 | 13.02 | 6.05 |
| Piauí | 2020-05-06 | 1032 | 1012 | 1053 | 1051 | 1.78 | - |
|  | 2020-05-07 | 1111 | 1074 | 1147 | 1131 | 1.81 | - |
|  | 2020-05-08 | 1192 | 1133 | 1250 | 1233 | 3.36 | - |
|  | 2020-05-09 | 1271 | 1188 | 1354 | 1278 | 0.54 | - |
|  | 2020-05-10 | 1351 | 1242 | 1461 | 1332 | 1.45 | - |
|  | 2020-05-11 | 1431 | 1292 | 1570 | 1443 | 0.82 | - |
| Rio Grande do Norte | 2020-05-06 | 1724 | 1654 | 1794 | 1739 | 0.86 | - |
|  | 2020-05-07 | 1804 | 1695 | 1913 | 1821 | 0.93 | - |
|  | 2020-05-08 | 1884 | 1737 | 2031 | 1919 | 1.82 | - |
|  | 2020-05-09 | 1964 | 1779 | 2149 | 1919 | 2.35 | - |
|  | 2020-05-10 | 2044 | 1819 | 2269 | 1919 | 6.52 | - |
|  | 2020-05-11 | 2124 | 1858 | 2390 | 1989 | 6.79 | - |
| Sergipe | 2020-05-06 | 977 | 938 | 1016 | 998 | 2.08 | - |
|  | 2020-05-07 | 1055 | 999 | 1111 | 1214 | 13.12 | 8.52 |
|  | 2020-05-08 | 1171 | 1083 | 1260 | 1438 | 18.55 | 12.39 |
|  | 2020-05-09 | 1236 | 1108 | 1363 | 1588 | 22.18 | 14.16 |
|  | 2020-05-10 | 1341 | 1178 | 1504 | 1771 | 24.29 | 15.09 |
|  | 2020-05-11 | 1431 | 1220 | 1642 | 1800 | 20.49 | 8.77 |

**Table 3:** Results of projections of confirmed cases of covid-19 between May 6 and 11, 2020 for the states of the Northern Region.

| state | date | predicted value | lower limit | upper limit | actual cases | absolute percentage error* | absolute percentage error** |
| --- | --- | --- | --- | --- | --- | --- | --- |
| Acre | 2020-05-06 | 889 | 870 | 908 | 943 | 5.71 | 3.69 |
|  | 2020-05-07 | 968 | 931 | 1005 | 1014 | 4.54 | 0.90 |
|  | 2020-05-08 | 1041 | 977 | 1105 | 1177 | 11.56 | 6.13 |
|  | 2020-05-09 | 1118 | 1025 | 1211 | 1335 | 16.26 | 9.27 |
|  | 2020-05-10 | 1192 | 1064 | 1320 | 1447 | 17.63 | 8.77 |
|  | 2020-05-11 | 1268 | 1102 | 1434 | 1460 | 13.16 | 1.81 |
| Amazonas | 2020-05-06 | 8833 | 8656 | 9010 | 9243 | 4.43 | 2.52 |
|  | 2020-05-07 | 9557 | 9218 | 9897 | 10099 | 5.36 | 2.00 |
|  | 2020-05-08 | 10282 | 9754 | 10809 | 10727 | 4.15 | - |
|  | 2020-05-09 | 11006 | 10267 | 11744 | 11925 | 7.71 | 1.52 |
|  | 2020-05-10 | 11730 | 10759 | 12701 | 12599 | 6.90 | - |
|  | 2020-05-11 | 12454 | 11231 | 13677 | 12919 | 3.60 | - |
| Amapá | 2020-05-06 | 2140 | 2052 | 2227 | 2046 | 4.58 | - |
|  | 2020-05-07 | 2348 | 2189 | 2508 | 2199 | 6.79 | - |
|  | 2020-05-08 | 2557 | 2316 | 2798 | 2322 | 10.11 | - |
|  | 2020-05-09 | 2765 | 2435 | 3096 | 2493 | 10.93 | - |
|  | 2020-05-10 | 2974 | 2545 | 3403 | 2613 | 13.82 | - |
|  | 2020-05-11 | 3183 | 2647 | 3718 | 2671 | 19.16 | - |
| Pará | 2020-05-06 | 5136 | 4991 | 5281 | 5524 | 7.03 | 4.40 |
|  | 2020-05-07 | 5516 | 5267 | 5765 | 5935 | 7.06 | 2.87 |
|  | 2020-05-08 | 5896 | 5536 | 6256 | 6519 | 9.56 | 4.04 |
|  | 2020-05-09 | 6276 | 5796 | 6756 | 7018 | 10.58 | 3.74 |
|  | 2020-05-10 | 6656 | 6046 | 7265 | 7348 | 9.42 | 1.13 |
|  | 2020-05-11 | 7035 | 6288 | 7783 | 8069 | 12.81 | 3.54 |
| Rondônia | 2020-05-06 | 966 | 937 | 995 | 943 | 2.44 | - |
|  | 2020-05-07 | 1071 | 1007 | 1135 | 1098 | 2.46 | - |
|  | 2020-05-08 | 1176 | 1068 | 1284 | 1222 | 3.76 | - |
|  | 2020-05-09 | 1281 | 1123 | 1439 | 1263 | 1.43 | - |
|  | 2020-05-10 | 1386 | 1172 | 1600 | 1302 | 6.45 | - |
|  | 2020-05-11 | 1491 | 1216 | 1766 | 1398 | 6.65 | - |
| Roraima | 2020-05-06 | 1160 | 1072 | 1248 | 932 | 24.46 | 15.06 |
|  | 2020-05-07 | 1451 | 1255 | 1647 | 1020 | 42.25 | 23.04 |
|  | 2020-05-08 | 1742 | 1414 | 2070 | 1124 | 54.98 | 25.81 |
|  | 2020-05-09 | 2033 | 1553 | 2513 | 1202 | 69.13 | 29.20 |
|  | 2020-05-10 | 2324 | 1674 | 2974 | 1290 | 80.16 | 29.77 |
|  | 2020-05-11 | 2615 | 1779 | 3451 | 1295 | 101.93 | 37.37 |
| Tocantins | 2020-05-06 | 381 | 370 | 392 | 423 | 9.90 | 7.24 |
|  | 2020-05-07 | 420 | 405 | 434 | 494 | 15.00 | 12.07 |
|  | 2020-05-08 | 459 | 437 | 481 | 572 | 19.81 | 15.97 |
|  | 2020-05-09 | 497 | 465 | 530 | 688 | 27.70 | 22.97 |
|  | 2020-05-10 | 536 | 491 | 581 | 747 | 28.21 | 22.17 |
|  | 2020-05-11 | 575 | 516 | 634 | 828 | 30.55 | 23.37 |

**Table 4:** Results of projections of confirmed cases of Covid-19 between May 6 and 11, 2020 for the states of the Midwest Region of Brazil and the Distrito Federal.

| state | date | predicted value | lower limit | upper limit | actual cases | absolute percentage error* | absolute percentage error** |
| --- | --- | --- | --- | --- | --- | --- | --- |
| Distrito Federal | 2020-05-06 | 1910 | 1868 | 1951 | 2046 | 6.66 | 4.63 |
|  | 2020-05-07 | 1982 | 1914 | 2051 | 2258 | 12.21 | 9.17 |
|  | 2020-05-08 | 2055 | 1959 | 2151 | 2442 | 15.85 | 11.90 |
|  | 2020-05-09 | 2128 | 2002 | 2253 | 2576 | 17.40 | 12.52 |
|  | 2020-05-10 | 2200 | 2044 | 2357 | 2682 | 17.96 | 12.11 |
|  | 2020-05-11 | 2273 | 2083 | 2463 | 2799 | 18.79 | 12.01 |
| Goiás | 2020-05-06 | 958 | 931 | 986 | 1024 | 6.43 | 3.76 |
|  | 2020-05-07 | 994 | 951 | 1038 | 1027 | 3.18 | - |
|  | 2020-05-08 | 1031 | 971 | 1090 | 1053 | 2.13 | - |
|  | 2020-05-09 | 1067 | 991 | 1143 | 1069 | 0.21 | - |
|  | 2020-05-10 | 1103 | 1010 | 1196 | 1093 | 0.91 | - |
|  | 2020-05-11 | 1139 | 1028 | 1250 | 1100 | 3.56 | - |
| Mato Grosso do Sul | 2020-05-06 | 290 | 280 | 300 | 288 | 0.62 | - |
|  | 2020-05-07 | 297 | 282 | 311 | 311 | 4.64 | - |
|  | 2020-05-08 | 303 | 284 | 323 | 326 | 6.95 | 1.00 |
|  | 2020-05-09 | 310 | 286 | 334 | 346 | 10.37 | 3.50 |
|  | 2020-05-10 | 317 | 289 | 345 | 362 | 12.46 | 4.69 |
|  | 2020-05-11 | 324 | 291 | 356 | 385 | 15.93 | 7.48 |
| Mato Grosso | 2020-05-06 | 378 | 365 | 392 | 385 | 1.74 | - |
|  | 2020-05-07 | 391 | 370 | 411 | 419 | 6.78 | 1.80 |
|  | 2020-05-08 | 403 | 375 | 430 | 464 | 13.17 | 7.26 |
|  | 2020-05-09 | 415 | 381 | 449 | 502 | 17.29 | 10.55 |
|  | 2020-05-10 | 427 | 387 | 468 | 519 | 17.63 | 9.87 |
|  | 2020-05-11 | 440 | 393 | 487 | 545 | 19.31 | 10.70 |

**Table 5:** Results of projections of confirmed cases of Covid-19 between May 6 and 11, 2020 for the states of the Midwest Region and the Distrito Federal.

| state | date | predicted value | lower limit | upper limit | actual cases | absolute percentage error* | absolute percentage error** |
| --- | --- | --- | --- | --- | --- | --- | --- |
| Espírito Santo | 2020-05-06 | 3710 | 3607 | 3814 | 3714 | 0.10 | - |
|  | 2020-05-07 | 3880 | 3709 | 4052 | 3988 | 2.70 | - |
|  | 2020-05-08 | 4050 | 3809 | 4291 | 4242 | 4.52 | - |
|  | 2020-05-09 | 4221 | 3906 | 4535 | 4412 | 4.34 | - |
|  | 2020-05-10 | 4391 | 3998 | 4784 | 4599 | 4.53 | - |
|  | 2020-05-11 | 4561 | 4085 | 5037 | 4819 | 5.36 | - |
| Minas Gerais | 2020-05-06 | 2596 | 2547 | 2646 | 2605 | 0.34 | - |
|  | 2020-05-07 | 2760 | 2691 | 2830 | 2770 | 0.35 | - |
|  | 2020-05-08 | 2938 | 2845 | 3031 | 2943 | 0.17 | - |
|  | 2020-05-09 | 3124 | 3000 | 3248 | 3123 | 0.03 | - |
|  | 2020-05-10 | 3315 | 3150 | 3481 | 3237 | 2.43 | - |
|  | 2020-05-11 | 3511 | 3295 | 3727 | 3320 | 5.74 | - |
| Rio de Janeiro | 2020-05-06 | 12911 | 12697 | 13124 | 13295 | 2.89 | - |
|  | 2020-05-07 | 13506 | 13230 | 13782 | 14156 | 4.59 | 2.64 |
|  | 2020-05-08 | 14101 | 13728 | 14474 | 15741 | 10.42 | 8.05 |
|  | 2020-05-09 | 14696 | 14200 | 15191 | 16929 | 13.19 | 10.26 |
|  | 2020-05-10 | 15291 | 14652 | 15929 | 17062 | 10.38 | 6.64 |
|  | 2020-05-11 | 15885 | 15088 | 16683 | 17939 | 11.45 | 7.00 |
| São Paulo | 2020-05-06 | 35672 | 34841 | 36503 | 37853 | 5.76 | 3.57 |
|  | 2020-05-07 | 37010 | 35483 | 38537 | 39928 | 7.31 | 3.48 |
|  | 2020-05-08 | 38348 | 36214 | 40482 | 41830 | 8.32 | 3.22 |
|  | 2020-05-09 | 39686 | 36958 | 42414 | 44411 | 10.64 | 4.50 |
|  | 2020-05-10 | 41024 | 37694 | 44353 | 45444 | 9.73 | 2.40 |
|  | 2020-05-11 | 42362 | 38416 | 46308 | 46131 | 8.17 | - |

**Table 6:** Results of projections of confirmed cases of Covid-19 between May 6 and 11, 2020 for the states of the Southern Region of Brazil.

| state | date | predicted cases | lower limit | upper limit | actual cases | absolute percentage error* | absolute percentage error** |
| --- | --- | --- | --- | --- | --- | --- | --- |
| Paraná | 2020-05-06 | 1643 | 1606 | 1680 | 1647 | 0.26 | - |
|  | 2020-05-07 | 1682 | 1616 | 1748 | 1678 | 0.23 | - |
|  | 2020-05-08 | 1721 | 1629 | 1813 | 1734 | 0.75 | - |
|  | 2020-05-09 | 1760 | 1642 | 1879 | 1809 | 2.70 | - |
|  | 2020-05-10 | 1799 | 1654 | 1944 | 1859 | 3.21 | - |
|  | 2020-05-11 | 1838 | 1666 | 2011 | 1873 | 1.85 | - |
| Santa Catarina | 2020-05-06 | 2911 | 2716 | 3107 | 2917 | 0.19 | - |
|  | 2020-05-07 | 3028 | 2735 | 3321 | 3082 | 1.75 | - |
|  | 2020-05-08 | 3144 | 2764 | 3524 | 3205 | 1.89 | - |
|  | 2020-05-09 | 3261 | 2797 | 3724 | 3372 | 3.30 | - |
|  | 2020-05-10 | 3377 | 2831 | 3923 | 3429 | 1.51 | - |
|  | 2020-05-11 | 3494 | 2865 | 4123 | 3529 | 1.00 | - |
| Rio Grande do Sul | 2020-05-06 | 2167 | 2109 | 2224 | 2100 | 3.17 | - |
|  | 2020-05-07 | 2233 | 2138 | 2328 | 2182 | 2.35 | - |
|  | 2020-05-08 | 2283 | 2166 | 2401 | 2493 | 8.41 | 3.71 |
|  | 2020-05-09 | 2367 | 2233 | 2500 | 2542 | 6.90 | 1.66 |
|  | 2020-05-10 | 2492 | 2340 | 2645 | 2576 | 3.25 | - |
|  | 2020-05-11 | 2640 | 2458 | 2821 | 2808 | 6.00 | - |

The results of the Table 2 show the results of the ARIMA models for the Northeast Region of Brazil. For the states of Bahia, Maranhão, Piauí and Rio Grande do Norte, the actual cases were within the range between the forecast limits from 06 to 11 May. The ARIMA model for the State of Piauí showed the best performance. The errors obtained, between the real cases and the estimated cases, varied between 0.54% to 3.36%. On the other hand, the model that had the worst performance for this region was the State of Sergipe. For that state, from the six days of forecast, only the cases estimated for May 6 were within the forecast interval. On the remaining days, the real cases of the accumulated cases of Covid-19 exceeded the maximum limits of the projections, with errors (between the real cases and the maximum limits) of 8.52%, 12.39%, 14.16%, 15.09% and 8.77%.

Table 3 shows the forecasts results for all states in the Brazilian Northern Region. The models that presented the best performance were the models for the states of Amapá and Rondônia. For the six days of forecast the actual cases were within the range between the minimum and maximum limits. The worst performances were for the state of Roraima and Tocantins. For those states, the errors between the actual cases and the minimum and maximum limits, respectively, reached more than 55% and 25%.

The forecasts results for the states of the Midwest Region and the Distrito Federal are presented in Table 4. The models referring to the states of Mato Grosso do Sul and Goiás, presented the best performances for the region. In the case of the State of Goiás, only the prediction for May 6, was not within the forecast interval. However, the absolute percentage error between the actual cases and the upper limit, did not exceed the 5% mark. In contrast, for the State of Mato Grosso do Sul, the cumulative sum of the actual cases of Covid19 was not within the forecast interval. However, the errors considering the actual cases and the upper limit, were not greater than 5%. On the other hand, both the model for the state of Mato Grosso and the model for the Distrito Federal presented a low performance. The last one being the one which showed the worst model for the region. In which, from the 6 days of forecast, all of them were outside the interval between the lower limit and the upper limit. Additionaly, only the cumulative sum of the actual cases for May 6 obtained an error below 5% in relation to the upper limit.

The results of the predictions for the states of the Southeast Region are presented in the Table 5. Among the four models generated, only the model from the State of Rio de Janeiro did not show a good performance. From the six days of prediction, only two met the criteria of a good performace. For the states of Espírito Santo and Minas Gerais, the forescasts for the six days were within the forecast interval. In these two states, the absolute percentage errors between the actual cases and the predicted values varied between 0.10% and 5.36%, for the State of Espírito Santo, and between 0.03% and 5.74%, for the State of Minas Gerais. The actual cases for the State of São Paulo, for most of the forecast days, were outside the lower limit and upper limit interval. However, for those days (May 6th to 10th), the errors between the actual cases and the upper limits ranged from 2.40% to 3.57%.

The ARIMA models generated for the South Region, as shown in the Table 6, obtained good results for the forecasts of the cumulative sum of Covid-19 cases for this region. For both the State of Santa Catarina and the State of Paraná, the actual cases were within the prediction interval and very close to the estimated values. The absolute percentage errors between the real cases of the Covid-19 accumulated cases for the two states varied between 0.26% and 3.21%, and 0.19% and 3.30%, respectively. In the case of the forecasts for Rio Grande do Sul, the actual cases of May 8 and 9 are outside the prediction interval range, however the errors between the actual case number and the upper limit were 3.17% and 1.66%, respectively.

The results of the forecasts for Brazil are shown in the Table 7. From May 6th to 10th, the actual cases were outside the prediction range (they were higher than the upper limit). However, the absolute percentage error between the upper limit and the actual cases did not exceed the 5% limit. On May 11, the value of the actual cases was within the prediction interval, showing an error of 5.63% between the predicted value and the actual cases.

**Table 7:**
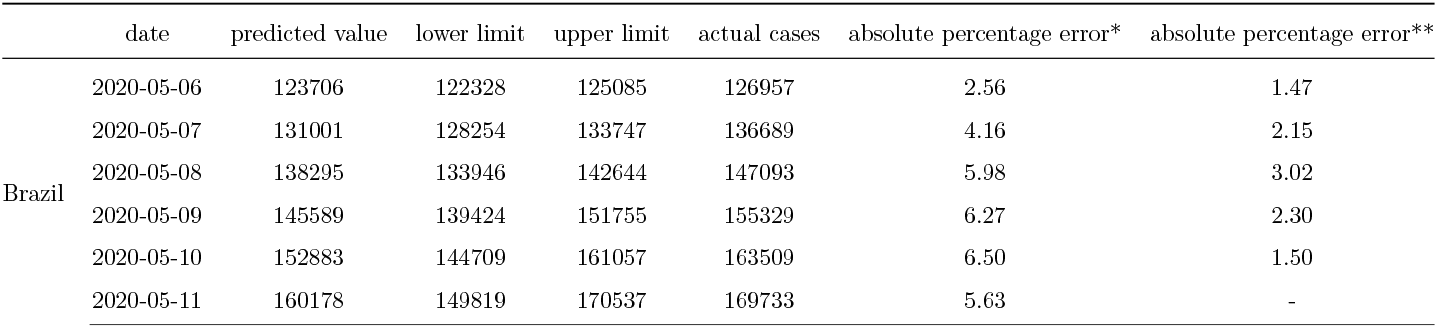
Result of the projections of the accumulated confirmed cases of Covid-19 for Brazil between May 6th to 11th, 2020.

### 4.2. Web application

The prototype of the developed system can be accessed by the link (https://www.cin.ufpe.br/~covidsgis). On the home screen, it is possible to visualize information about spatial predictions (Figure 10). The projections of the cumulative cases of Covid-19 can be accessed in the “More information” option, in which the user will be directed to the page with the graphics. In this screen, graphs of the temporal predictions, (Figure 10), as well as graphs referring to the distribution of daily cases and deaths in Brazil (Figure 12) are also available. In addition, it is available daily and cumulative cases (13), as well as daily and cumulative deaths (14). For these graphs, the user can select from which state they wish to evaluate these information. The software’s back-end is freely available for non-commercial purpose on our Github repository: https://github.com/Biomedical-Computing-UFPE/Covid-SGIS.

## 5. Discussion

In this study, we evaluated the forecast of the cumulative cases of Covid-19 for Brazil, and its 27 federative units in Brazil, using ARIMA models in the period between May 6 and 11. The results showed that, in general, for Brazil, the cases of Covid-19 were still on the rise during the observed period, as shown in the Figure 9. It shows the predictions of the accumulated cases of the disease from 06 to 11 of May to 2020. This rise can be confirmed by analizing the actual cases of Covid-19, for the same time period, in Table 7 which shows in detail the growth of the number of cases. In addition, it is not possible to observe that Brazil has reached the peak of the disease in this period, which was confirmed in the results presented in the Table 7.

Figures 5, 6, 7 and 8 show the forecasts curves for each of the states showed that the cases of Covid-19, in the observed period, were tending to increase. This fact was confirmed with the results in the Section 4. Both for Brazil and its federative units, the ARIMA models (p, d, q) managed to capture the trends in the curves. This can be useful for health managers and governments in promoting public policies to combat Covid-19.

**Figure 5:**
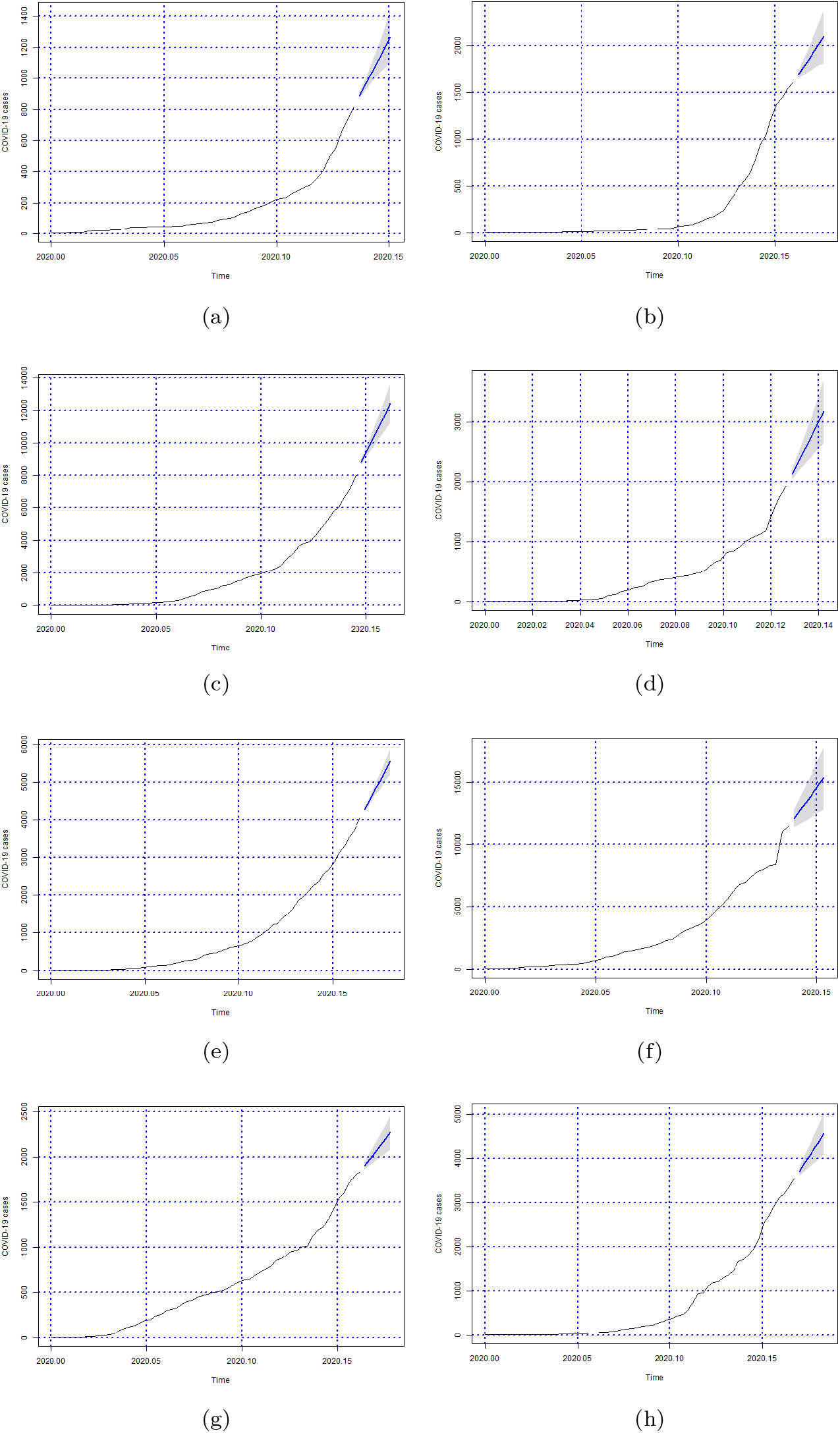
Forecasts of the number of Covid-19 cases from 06-05-2020 to 11-05-2020 for states (a) Acre, (b) Alagoas, (c) Amazonas, (d) Amapá, (e) Bahia, (f) Ceará, (g) Distrito Federal and (h) Espírito Santo.

**Figure 6:**
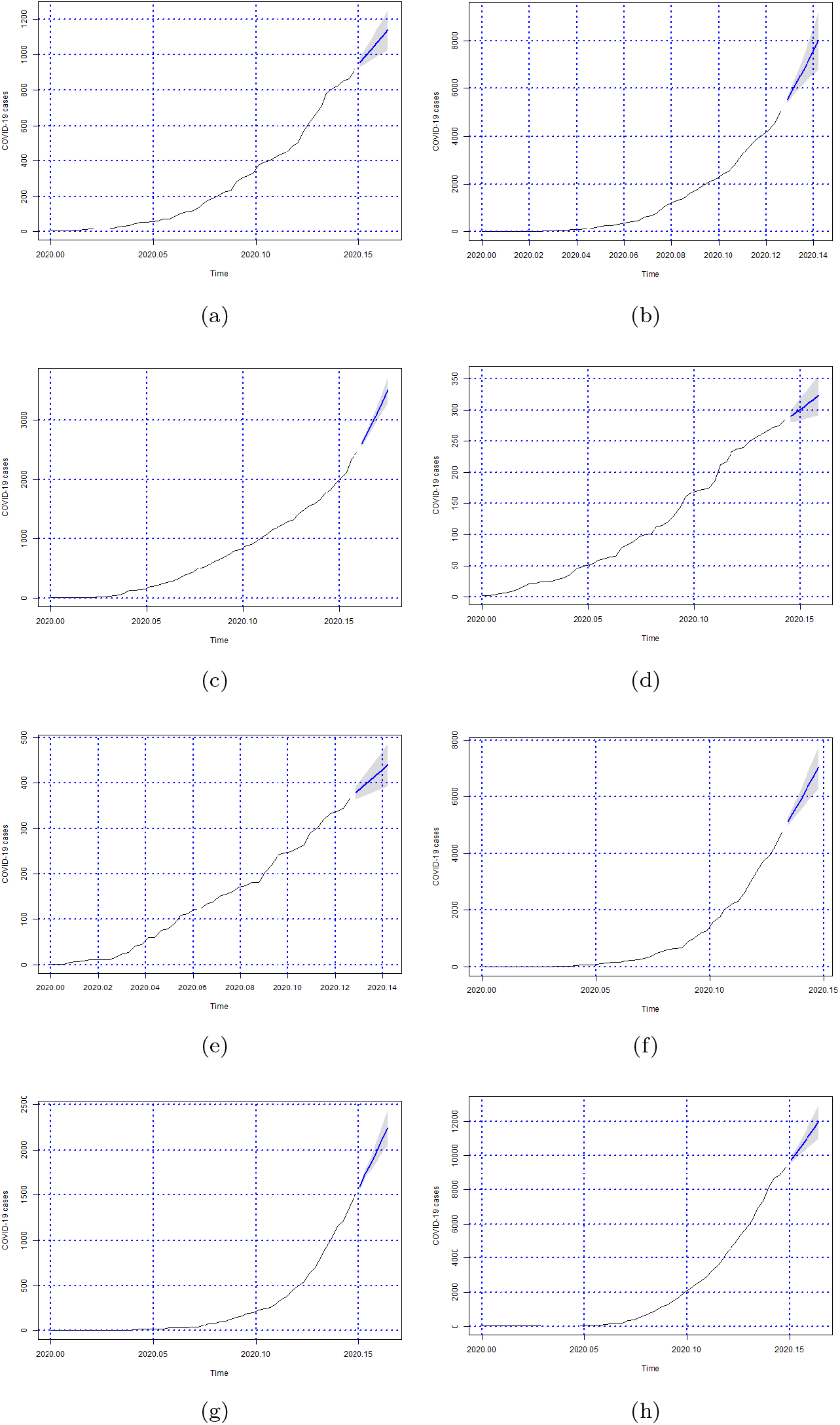
Forecasts of the number of Covid-19 cases from 06-05-2020 to 11-05-2020 for states (a) Goiás, (b) Maranhão, (c) Minas Gerais, (d) Mato Grosso do Sul, (e) Mato Grosso, (f) Pará, (g) Paraíba and (h) Pernambuco.

**Figure 7:**
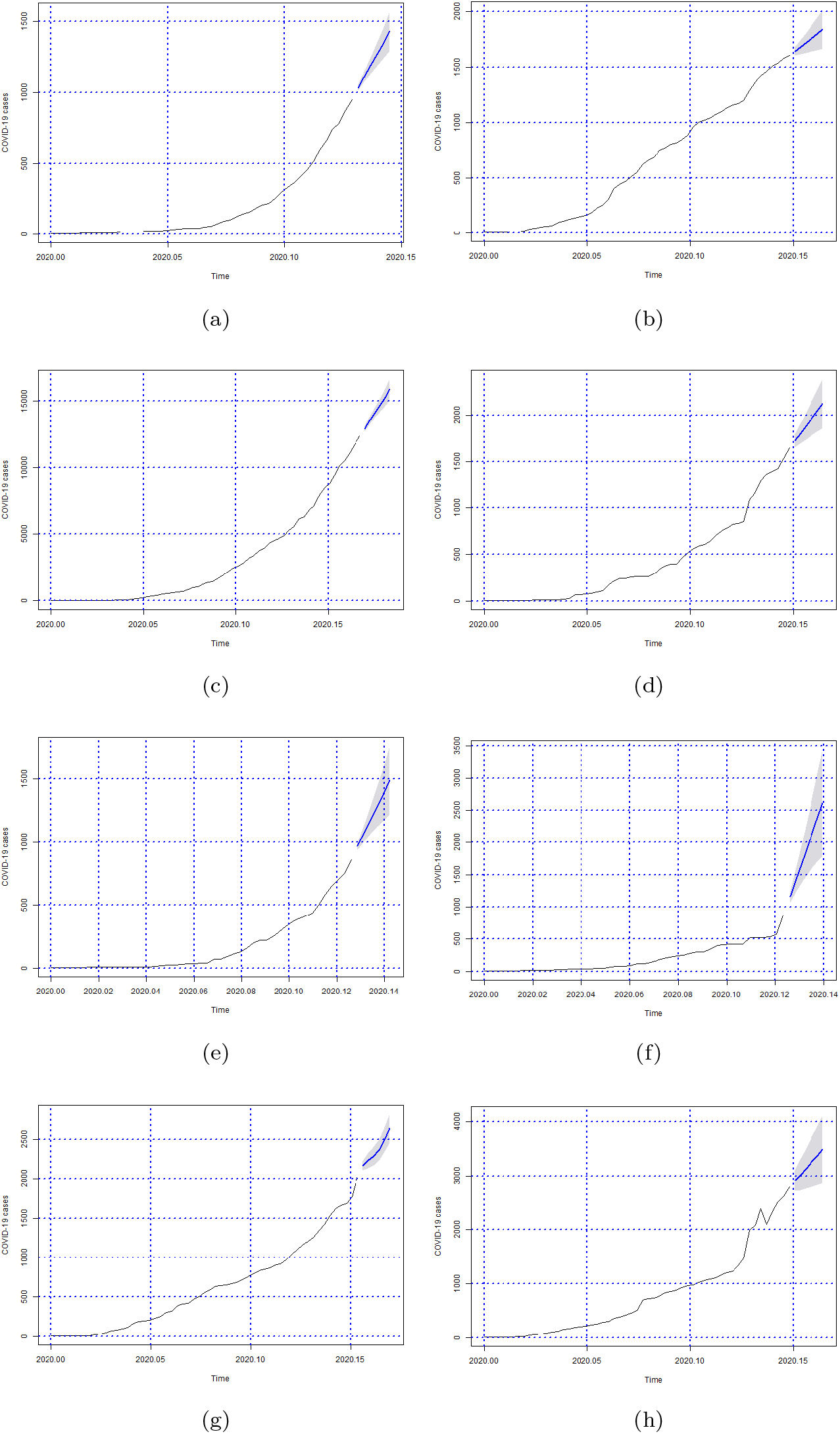
Forecasts of the number of Covid-19 cases from 06-05-2020 to 11-05-2020 for states (a) Piauí, (b) Paraná, (c) Rio de Janeiro, (d) Rio Grande do Norte, (e) Rondônia, (f) Roraima, (g) Rio Grande do Sul and (h) Santa Catarina.

**Figure 8:**
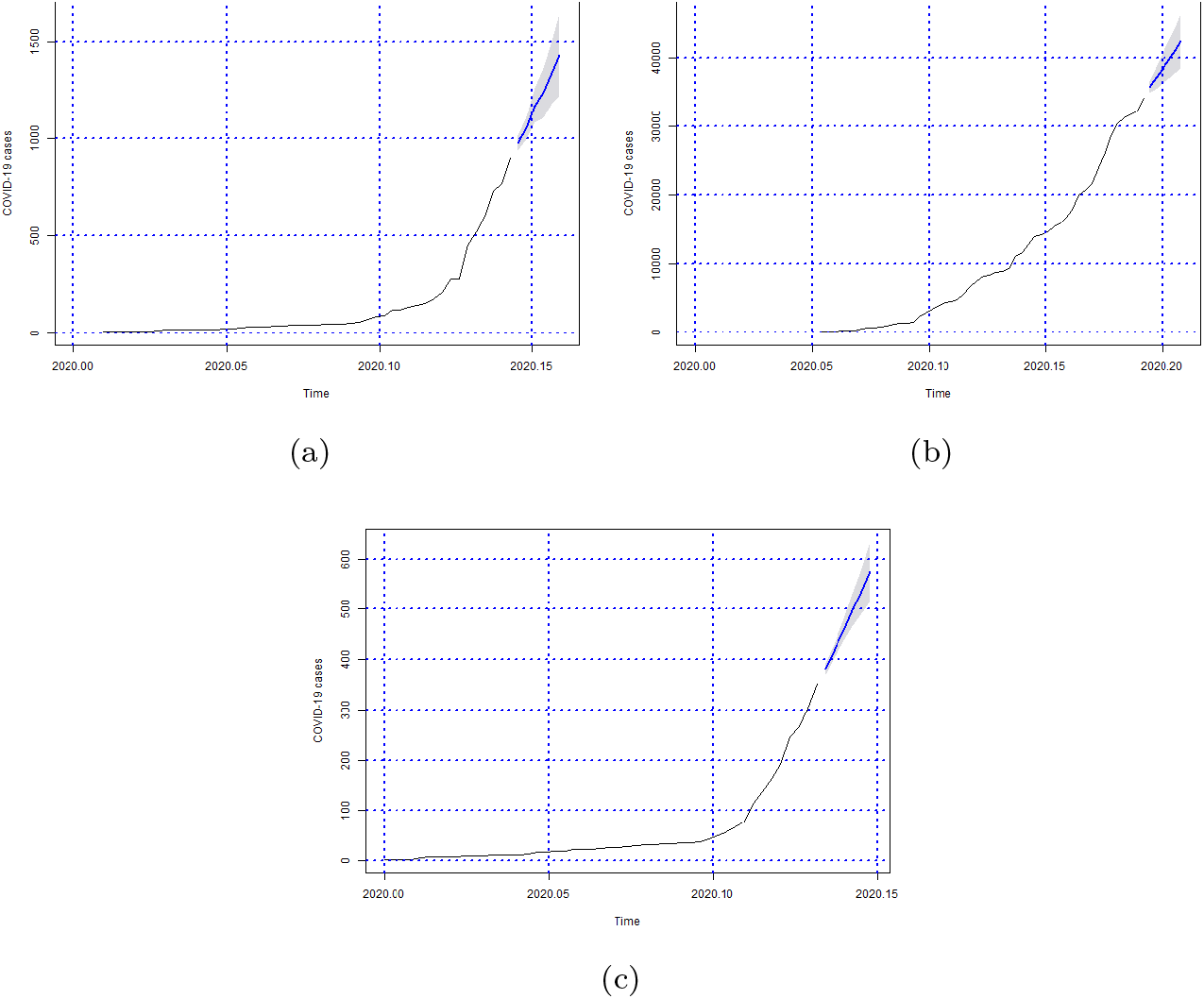
Forecasts of the number of Covid-19 cases from 06-05-2020 to 11-05-2020 for states (a) Sergipe, (b) São Paulo and (c) Tocantins.

**Figure 9:**
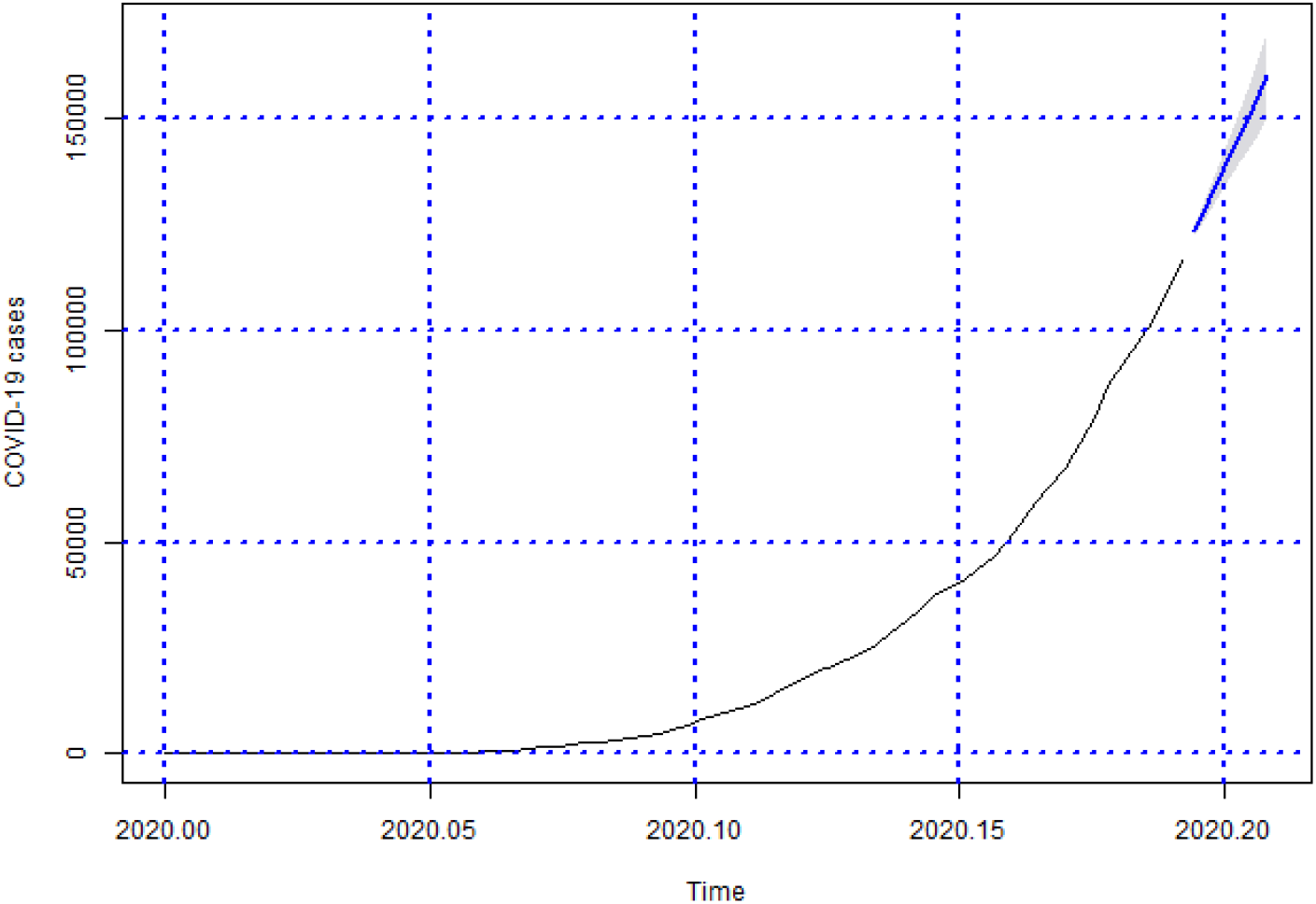
Forecasts of the number of Covid-19 cases from 06-05-2020 to 11-05-2020 for Brazil

**Figure 10:**
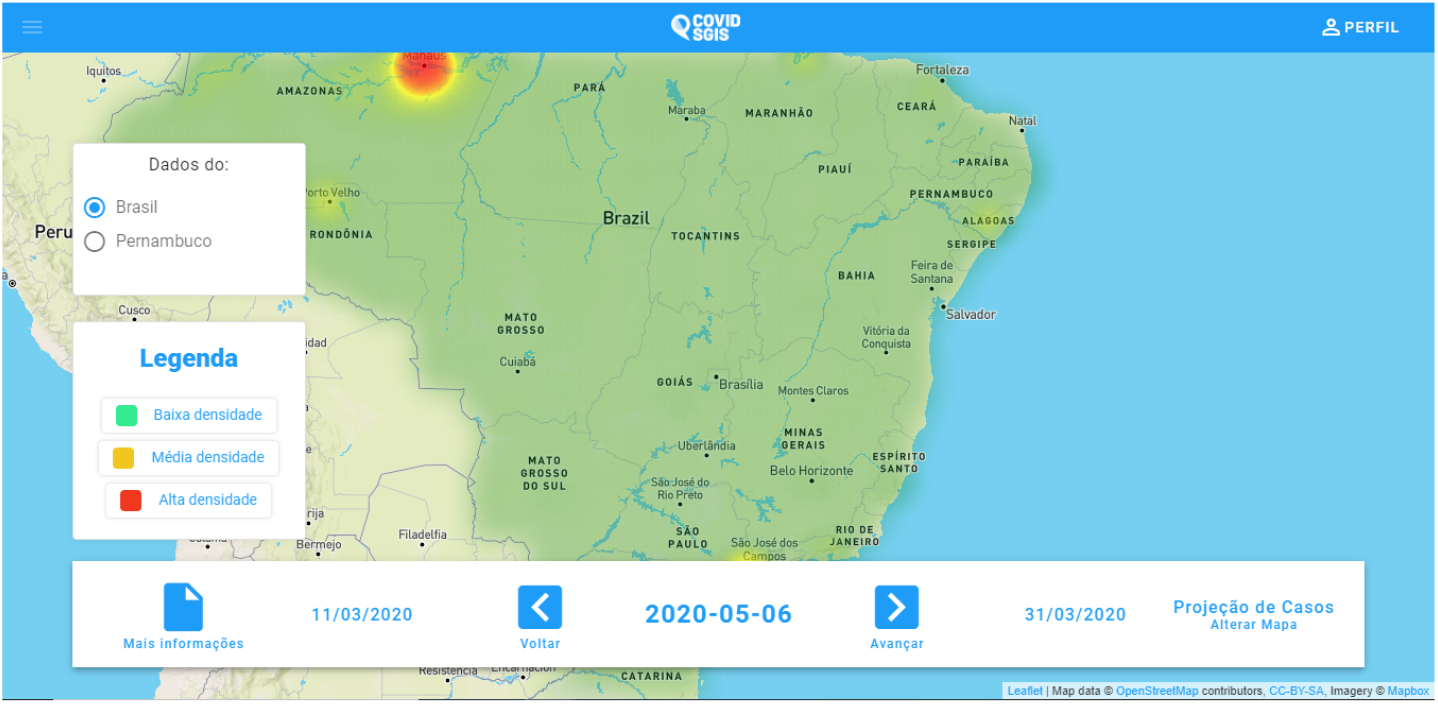
COVID SGIS web application home screen

**Figure 11:**
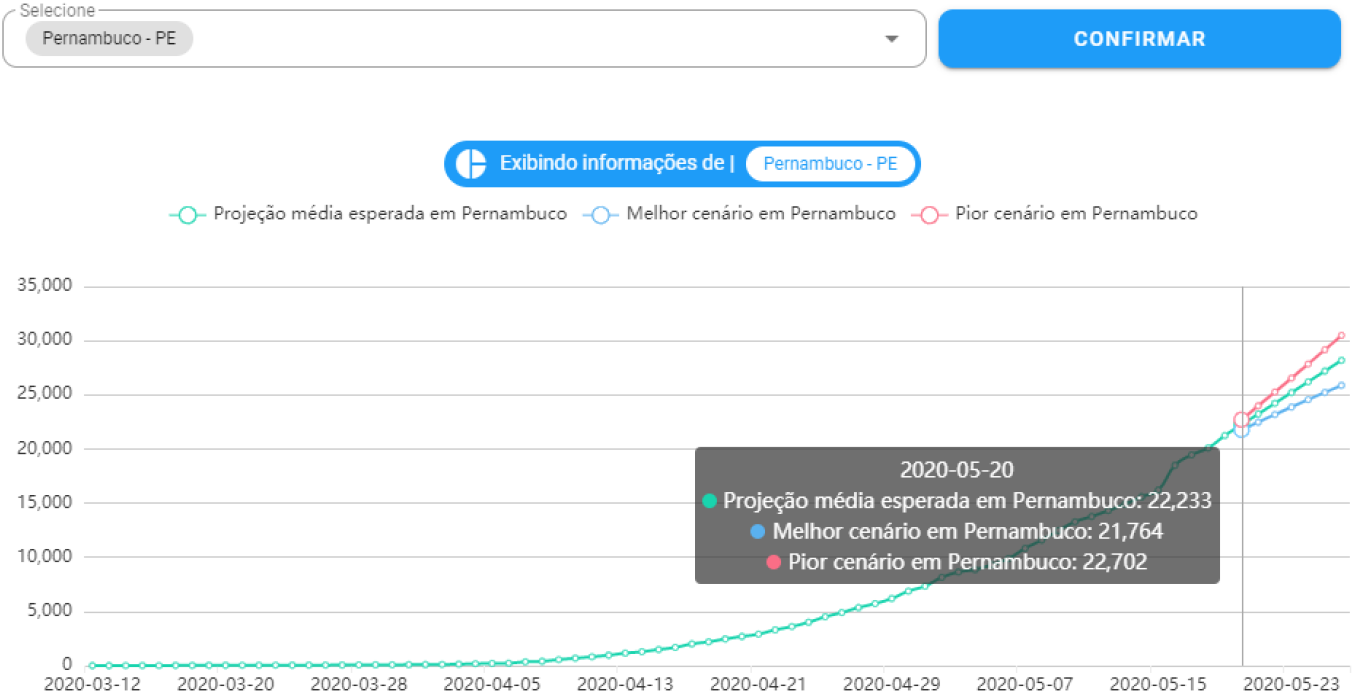
Accumulated cases of Covid-19 forecast graph. The prediction with ARIMA is represented by the green line. The worst case scenario (indicated by the upper limit of the prediction) is represented by the line in red. The best scenario (indicated by the lower limit of the prediction) is represented by the blue line.

**Figure 12:**
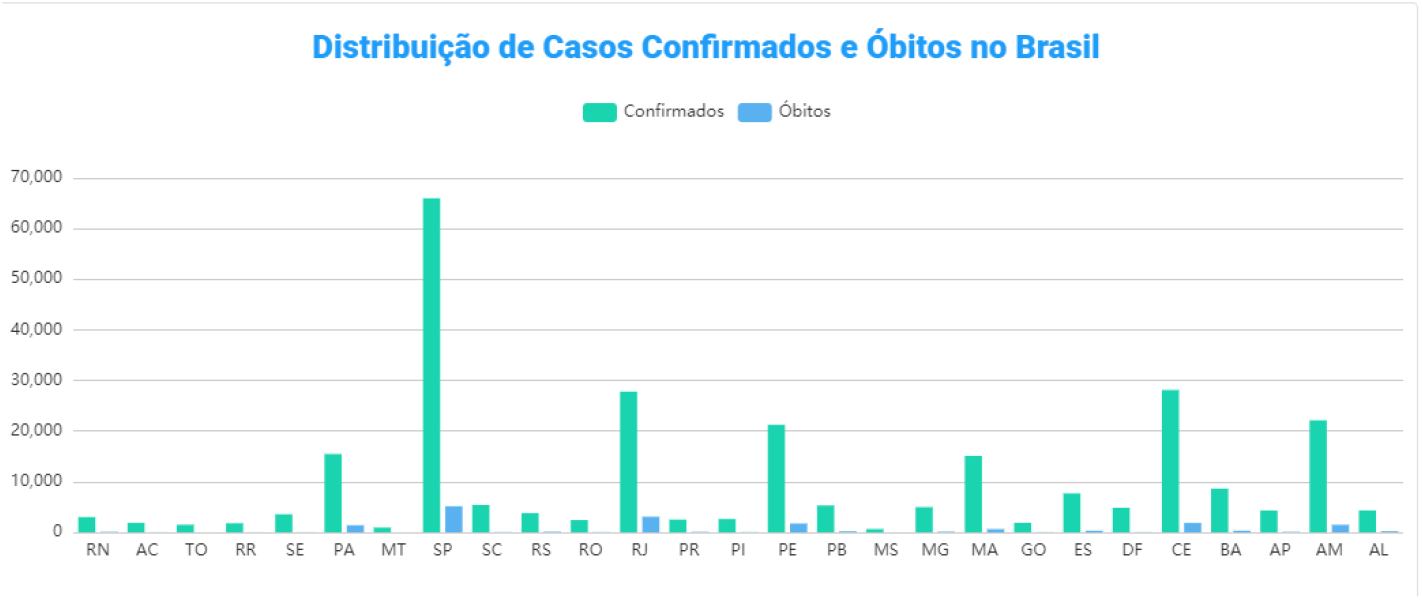
Screen of the graph of the distribution of confirmed cases and deaths by Covid-19. In this graph the user can have an overview of the accumulated confirmed cases and death cases in all states of Brazil and the Distrito Federal.

**Figure 13:**
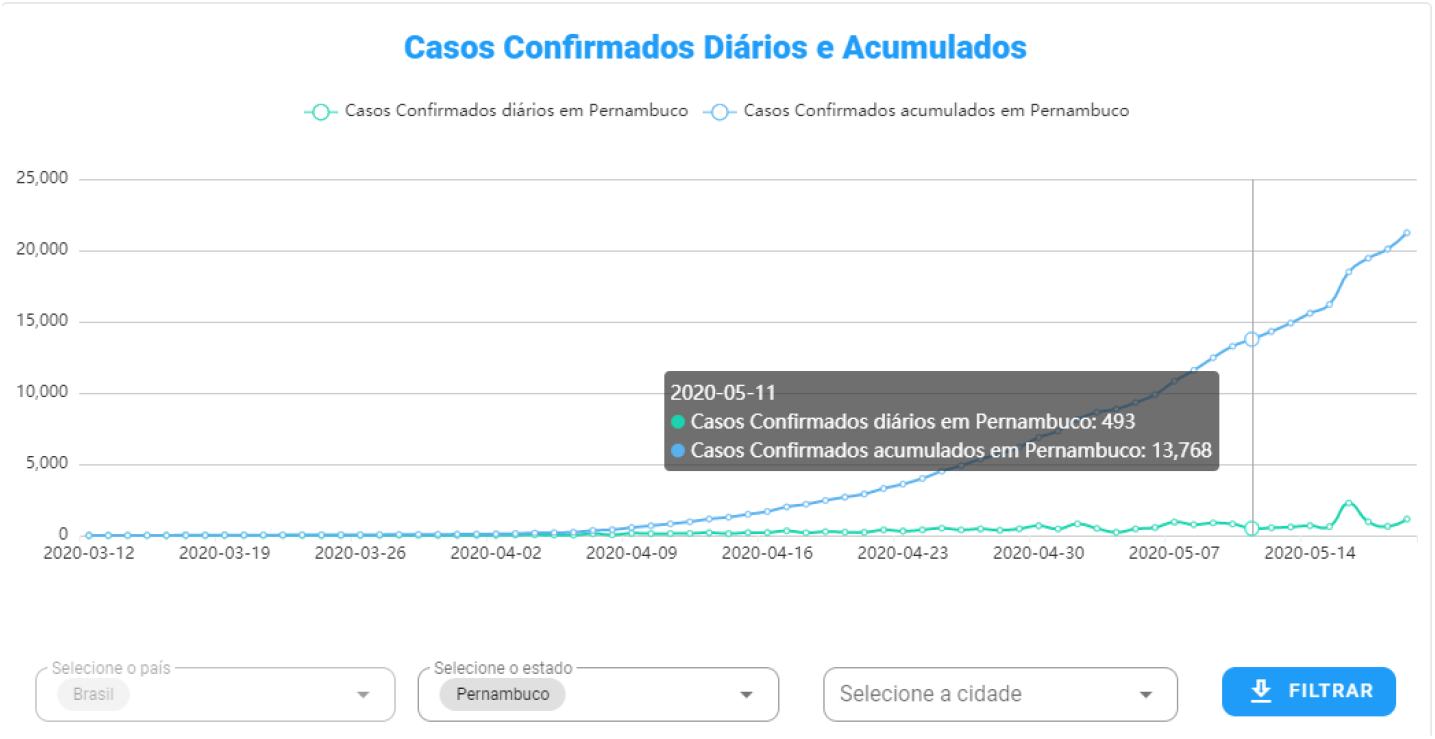
In COVID SGIS, the user can follow the daily and accumulated confirmed cases of Covid-19 for each Brazilian state and the Distrito Federal, separately.

**Figure 14:**
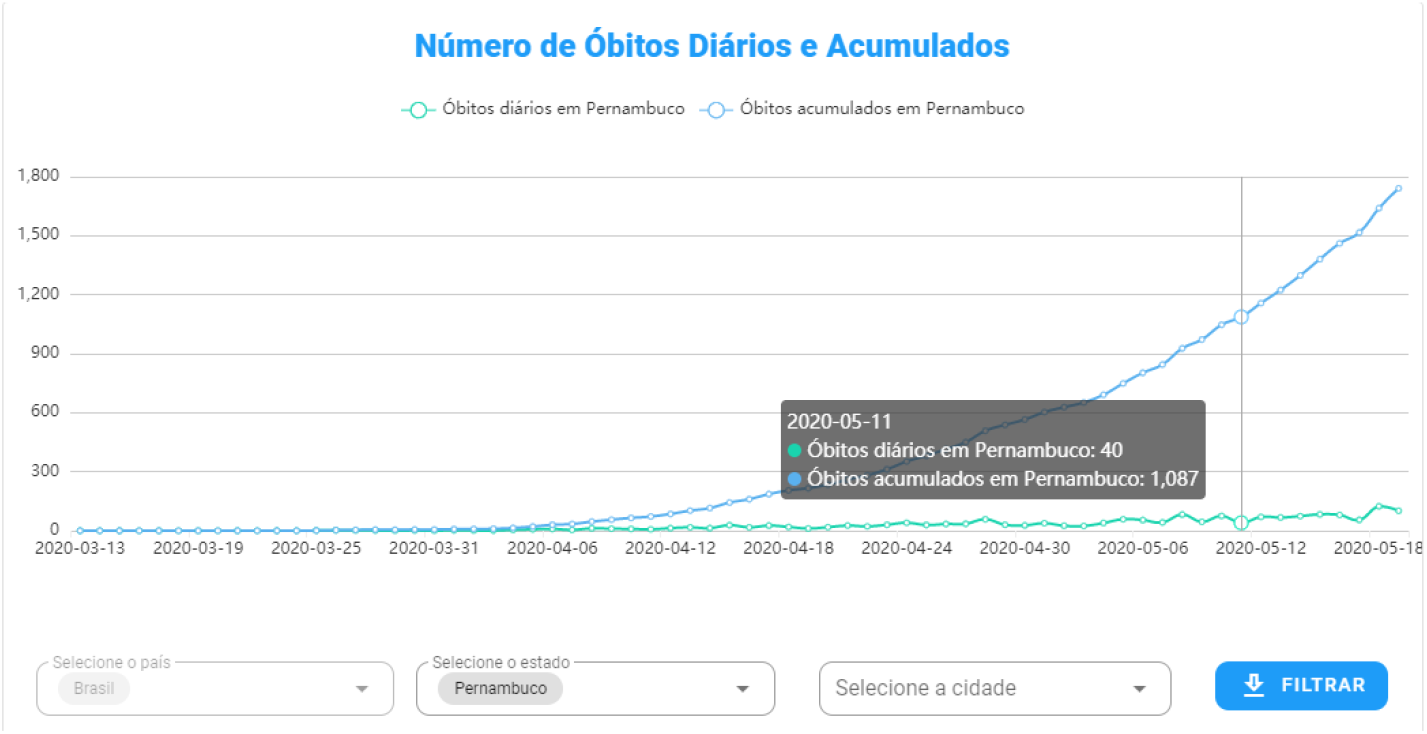
In COVID SGIS, the user can follow the daily and accumulated confirmed deaths from Covid-19 for each Brazilian state and the Distrito Federal, separately.

The low performance of the predictions for some Brazilian states may be associated with the underreporting of cases. That is, the amount of tests for the diagnosis is not sufficient to meet the populational demand. Thus, people who are asymptomatic or have symptoms of the disease, but have not been tested, are not counted in the reported cases. This problem makes the data substantial underestimated.

Another factor that may be associated with higher errors in the predictions is the time taken to update the databases from the Departments of Health. This directly affects the historical series of the states’ accumulated cases. As the modeling using ARIMA, depends a lot on the historical series, this delay in updating the databases, directly affect the real-time forecast. In addition, high prediction errors can also be associated with the historical series itself. As the day of the first notification varies from state to state, the amount of data assessed by the model before making the prediction is also changed.

Besides that data in Brazil are probably substantial underestimates considering that Federal Government insists in sow fake news and discouraging the social isolation (Prado, 2020). Another import issue is the social inequality of the population. According to Prado (2020) 13 million Brazilians live in favelas where it is common to have more than three people per room and the acess to clean water is precarious.

Among the limitations of this study, we have the fact that, for the prediction, we only used the accumulated case history of Covid-19 for each of the 27 federative units. Therefore, factors related to probable virus mutations were disregarded, as well as demographic information and risk factors.

## 6. Conclusion

Several countries are being greatly affected by the Covid-19 cases. With a high number of infected people and public health systems operating at maximum capacity, the situation is becoming increasingly critical. For this reason, it is important to have a tool that performs the prediction of Covid-19 cases, able to aid government officials, health managers and general stakeholders to ellaborate health policies and plan direct interventions. Therefore, it is possible to make short-term decisions, develop public policies, and direct resources to health professionals and hospitals. In addition, the forecasting tool can also assist governments in controlling measures of social isolation and lockdown. The evaluation of these predictions allows to intensify the restrictions of social agglomerations, as well as to evaluate the effect of these measures in the contagion of the population.

With that in mind, our motivation is to provide a robust, flexible and fast forecasting method. Thus, we combined the ARIMA model for time series analysis with Artificial Intelligence techniques. ARIMA was a good choice, being efficient in predicting a disease like Covid-19, which has a rapid proliferation and changes the scenario daily. The method is capable of presenting the prediction if the context of the prediction moment remains constant, as well as presenting the best and worst scenarios, with a confidence interval of 95%.

The developed models achieved a good performance taking into account the percentage errors. Among the 26 Brazilian states analyzed plus Distrito Federal, the majority presented satisfactory results, with low errors. Some states such as Roraima, Tocantins and Distrito Federal showed higher errors. Some hypotheses can be raised for this: many cases of Covid-19 are not reported, as in cases of asymptomatic people or who remained in isolation at home; in addition, database updates can be slow, directly affecting the prediction of the following days. This fact becomes even worse when taking into account the measures of the Brazilian Ministry of Health to limit testing to severe cases attested by clinical diagnosis.

Besides that, a specific issue in Brazil is the lack of leadership and collaborative spirit on the part of the Federal Government. The absence of these characteristics fosters fear, insecurity, and misinformation among Brazilian citizens. In this sense, the inaccurate information about Covid-19 itself is aggravated by the huge amount of fake news spreading all over the country financed by the person of the national president himself and his supporters. Despite that, State and Municipal governments are struggling to maintain the social isolation and provide more health care to the population, frequenly assuming responsibilities of the central government.

Another fundamental challenge resides on the strong social inequality in Brazil. Many Brazilian citizens do not have a fixed income. Since the Federal Government is not being efficient to ensure their basic survival needs, these people need to keep working. Therefore, making it difficult to implement social isolation policies and, consequently, increasing the size of the susceptible population and the circulation of asymptomatic individuals.

Finally, our method provides a possibility of dynamic forecasting: the proposed model is retrained and adapted to the real scenario in a daily basis. The proposed model is trained everyday with a maximum window of three days, achieving low errors in most predictions, and acceptable errors (inside the confidence interval) in the worst scenarios. This forecasting window is dynamic, since it is chosen by automatic ARIMA models. Another advantage of our proposal is the use of multiple databases. In this way, several countries can benefit from this solution, adapting the model to their databases, incorporating dedicated web crawlers, for instance. Our system can be an important tool to guide the course of this pandemic.

## Data Availability

The software code (back-end) is available at GitHub. The complete software can be used online.

## 7. Acknowledgements

The authors are grateful to the Brazilian research agencies FACEPE, CAPES and CNPq, for the partial financial support of this research.

## Compliance with Ethical Standards

This study was funded by the Federal University of Pernambuco and the Brazilian research agencies FACEPE, CAPES and CNPq.

All procedures performed in studies involving human participants were in accordance with the ethical standards of the institutional and/or national research committee and with the 1964 Helsinki declaration and its later amendments or comparable ethical standards.

## Footnotes

1 https://brasil.io/home/

2 https://brasil.io/dataset/covid19/caso/

3 https://www.r-project.org/

